# Effectiveness of 2 and 3 mRNA COVID-19 Vaccines Doses against Omicron and Delta-Related Outpatient Illness among Adults, October 2021 – February 2022

**DOI:** 10.1101/2022.04.06.22273535

**Authors:** Sara S. Kim, Jessie R. Chung, H. Keipp Talbot, Carlos G. Grijalva, Karen J. Wernli, Erika Kiniry, Emily T. Martin, Arnold S. Monto, Edward A. Belongia, Huong Q. McLean, Manjusha Gaglani, Mufaddal Mamawala, Mary Patricia Nowalk, Krissy Moehling Geffel, Sara Y. Tartof, Ana Florea, Justin S. Lee, Mark W. Tenforde, Manish M. Patel, Brendan Flannery, Strain Surveillance and Emerging Variants Team

## Abstract

**Background:** We estimated SARS-CoV-2 Delta and Omicron-specific effectiveness of 2 and 3 mRNA COVID-19 vaccine doses in adults against symptomatic illness in US outpatient settings.

**Methods:** Between October 1, 2021, and February 12, 2022, research staff consented and enrolled eligible participants who had fever, cough, or loss of taste or smell and sought outpatient medical care or clinical SARS-CoV-2 testing within 10 days of illness onset. Using the test-negative design, we compared the odds of receiving 2 or 3 mRNA COVID-19 vaccine doses among SARS-CoV-2 cases versus controls using logistic regression. Regression models were adjusted for study site, age, onset week, and prior SARS-CoV-2 infection. Vaccine effectiveness (VE) was calculated as (1 – adjusted odds ratio) x 100%.

**Results:** Among 3847 participants included for analysis, 574 (32%) of 1775 tested positive for SARS-CoV-2 during the Delta predominant period and 1006 (56%) of 1794 participants tested positive during the Omicron predominant period. When Delta predominated, VE against symptomatic illness in outpatient settings was 63% (95% CI: 51% to 72%) among mRNA 2-dose recipients and 96% (95% CI: 93% to 98%) for 3-dose recipients. When Omicron predominated, VE was 21% (95% CI: -6% to 41%) among 2-dose recipients and 62% (95% CI: 48% to 72%) among 3-dose recipients.

**Conclusions:** In this adult population, 3 mRNA COVID-19 vaccine doses provided substantial protection against symptomatic illness in outpatient settings when the Omicron variant became the predominant cause of COVID-19 in the U.S. These findings support the recommendation for a 3^rd^ mRNA COVID-19 vaccine dose.

## Background

On November 29, 2021, the Centers for Disease Control and Prevention (CDC) recommended that all adults aged ≥18 years receive a 3^rd^ mRNA COVID-19 vaccine booster dose at least 6 months after completing a 2-dose primary series [1]. The 6-month interval recommendation was shortened to at least 5 months on January 4, 2022, for the Pfizer-BioNTech vaccine and on January 7, 2022, for the Moderna vaccine. These recommendations were released during the emergence of the Omicron variant, which was first detected in the United States (US) on December 1, 2021 [2]. Effectiveness of 2 mRNA vaccines doses against symptomatic illness or hospitalization due to infection with the Omicron variant has been lower compared to the Delta variant, with increased protection against both variants after receipt of a 3^rd^ dose [3-5]. However, data comparing 2- and 3-dose vaccine effectiveness (VE) against symptomatic COVID-19 in outpatient settings during periods when the Delta and Omicron variants predominated are limited, especially among COVID-19 cases identified through active surveillance where all enrolled participants with COVID-19-like illness (CLI) are tested for SARS-CoV-2.

Studies with active enrollment such as the US Flu Vaccine Effectiveness Network (US Flu VE Network) provide access to specimens for research purposes including whole genome sequencing and access to data not available in medical records including risk factors for SARS-CoV-2 infection [6]. To assess the impact of a 3^rd^ dose in the context of emerging variants with immune evasion [7] and potential waning immunity, we estimated variant-specific effectiveness of 2 versus 3 mRNA vaccine doses against symptomatic illness in outpatient settings. Additionally, we utilized virus sequencing data to define periods when Delta and Omicron variants each predominated.

## Methods

### Study Design and Population

This study was conducted within the US Flu VE Network, which consists of participating health systems in 7 states: California, Michigan, Pennsylvania, Tennessee, Texas, Washington, and Wisconsin. Between October 1, 2021, and February 12, 2022, research staff screened patients seeking outpatient medical care or SARS-CoV-2 clinical testing using a standard case definition for CLI [8]. Eligible participants had an acute onset of fever, cough, or loss of taste/smell with symptom duration of <10 days [8] and had a clinical or research respiratory specimen collected for SARS-CoV-2 molecular testing within 10 days of illness onset. Research staff consented and enrolled eligible participants, who may have sought in-person medical care for CLI, completed a telehealth visit, or sought SARS-CoV-2 testing. Enrolled participants completed surveys with standardized questions across all research sites at enrollment including questions about demographics, symptoms experienced for current illness, COVID-19 vaccination history, prior SARS-CoV-2 infection, general health status, chronic medical conditions (heart disease, lung disease, diabetes, cancer, liver or kidney disease, immune suppression, or high blood pressure), and high-risk SARS-CoV-2 exposures (healthcare worker with close patient contact; contact with another laboratory-confirmed SARS-CoV-2 case in the 14 days before illness onset; or household member with laboratory-confirmed SARS-CoV-2 or with symptoms consistent with CLI in the 14 days before illness onset). This activity was reviewed and approved by the CDC and each US Flu VE Network site’s Institutional Review Board.^1^

### SARS-CoV-2 Status

Participants were tested for SARS-CoV-2 by reverse-transcription polymerase chain reaction tests using respiratory specimens collected for clinical or research purposes. We classified participants with a positive SARS-CoV-2 result as cases. Participants who had discordant clinical and research results were categorized as a case if at least one of the results were positive. We classified participants with only negative SARS-CoV-2 results as controls.

In addition, SARS-CoV-2 virus variants from a subset of SARS-CoV-2 positive participants with onset dates between November 9, 2021, and January 9, 2022, were identified by whole genome sequencing. Research-collected SARS-CoV-2 positive respiratory specimens with cycle threshold values <30 and stored in appropriate transport medium were prepared for sequencing using the xGen SARS-CoV-2 library preparation kit (Integrated DNA Technologies, Inc., Coralville, IA). Libraries were sequenced on a NovaSeq instrument (Illumina Inc., San Diego, CA). A single consensus genome for each sample was generated. SARS-CoV-2 variants were determined using Pangolin version 3.1.20 [pangoLEARN 1.2.123, Scorpio 0.3.16] [9].

### COVID-19 Vaccination Status

COVID-19 vaccination status was verified using electronic medical records, immunization information systems, and vaccination record cards. Participants considered vaccinated with 2 doses were those who received 2 mRNA vaccine doses ≥14 days before illness onset (2-dose). To be considered for the 2-dose analyses, participants must have received doses ≥16 days apart for Pfizer-BioNTech vaccines and ≥23 days apart for Moderna vaccines. Participants considered vaccinated with 3 doses were those who received 3 mRNA vaccine doses, where the 3^rd^ dose was given ≥7 days before illness onset (3-dose) [4]. Participants who received a 3^rd^ dose before the recommended ≥150 days after the 2^nd^ dose were also considered 3-dose recipients but excluded from sensitivity analyses. Three-dose recipients included both immunocompromised participants who received a 3^rd^ dose as a primary series and otherwise healthy participants who received a 3^rd^ dose as a booster. Those who did not report vaccine receipt and had no documentation of an mRNA COVID-19 vaccination before illness onset were defined as unvaccinated. We excluded participants who self-reported COVID-19 vaccination but were missing verified documentation of doses received.

### Statistical Analyses

We limited analyses to adults aged ≥18 years. Using the test-negative design [10], we compared the odds of 2- or 3-dose mRNA COVID-19 vaccination among COVID-19 cases versus test-negative controls using logistic regression. VE was calculated as (1 – adjusted odds ratio) x 100%. Regression models were adjusted for variables identified *a priori* including study site, age, and illness onset week. Sex, race and ethnicity, illness onset to specimen collection interval, self-reported high-risk exposure, self-reported chronic medical condition, and self-reported prior SARS-CoV-2 infection were evaluated as model covariates using a change-in-estimate (≥5% change in odds ratio) forward stepwise approach. We also evaluated chronic medical condition as a potential effect modifier (p-value <0.05). In addition to covariates included *a priori*, prior SARS-CoV-2 infection was included in the final regression model.

We evaluated VE by variant, either sequence-confirmed variant or using time periods of predominant Delta (illness onset of October 1 – December 9, 2021) versus Omicron circulation (illness onset of December 20, 2021 – February 12, 2022) when variant was not confirmed by sequencing. These periods were selected based on the SARS-CoV-2 sequencing results on a subset of cases in the US Flu VE Network. Due to co-circulation of the Delta and Omicron variants between December 10 – 19, 2021, we excluded participants without sequenced viruses with onset dates during this period for variant-specific estimates. We also assessed potential waning immunity among 2-dose recipients by comparing VE of those who received their 2^nd^ dose 14-149 days versus ≥150 days prior to illness onset during each variant predominant period.

We conducted several subgroup analyses where 3-dose VE was stratified by self-reported high-risk exposure status, self-reported chronic medical condition, self-reported prior SARS-CoV-2 infection, days between illness onset and specimen collection date, and self-reported presence of fever with cough or shortness of breath during the Delta and Omicron predominant periods. Analyses by illness onset to specimen collection interval were performed to identify bias resulting from potential false negative SARS-CoV-2 test results among participants who presented for care or testing later than those presenting 0-2 days after illness onset [10]. Analyses by symptoms were performed to evaluate VE among persons with potentially more severe illness compared to those without fever paired with cough or shortness of breath, indicating more mild illness.

## Results

### Study Population

Between October 2021 and February 2022, US Flu VE Network sites enrolled 4448 eligible outpatients aged ≥18 years, among whom 601 were excluded due to receiving a non-mRNA vaccine (n=216), self-reporting vaccination history with no documentation available (n=145), receiving 1 mRNA COVID-19 vaccine dose (n=121), missing vaccine product information (n=55), missing SARS-CoV-2 testing information (n=34), or having an indeterminate vaccination status (n=30). Among 3847 included for analysis, 575 (32%) of 1775 participants tested SARS-CoV-2 positive during the Delta predominant period and 1006 (56%) of 1794 participants tested positive during the Omicron predominant period. There were 278 participants whose illness onset dates fell between the defined Delta and Omicron predominance periods. SARS-CoV-2 positivity reached over 50% during the 3^rd^ week of December and peaked at 64% during the 2^nd^ week of January (Figure 1).

**Figure 1:**
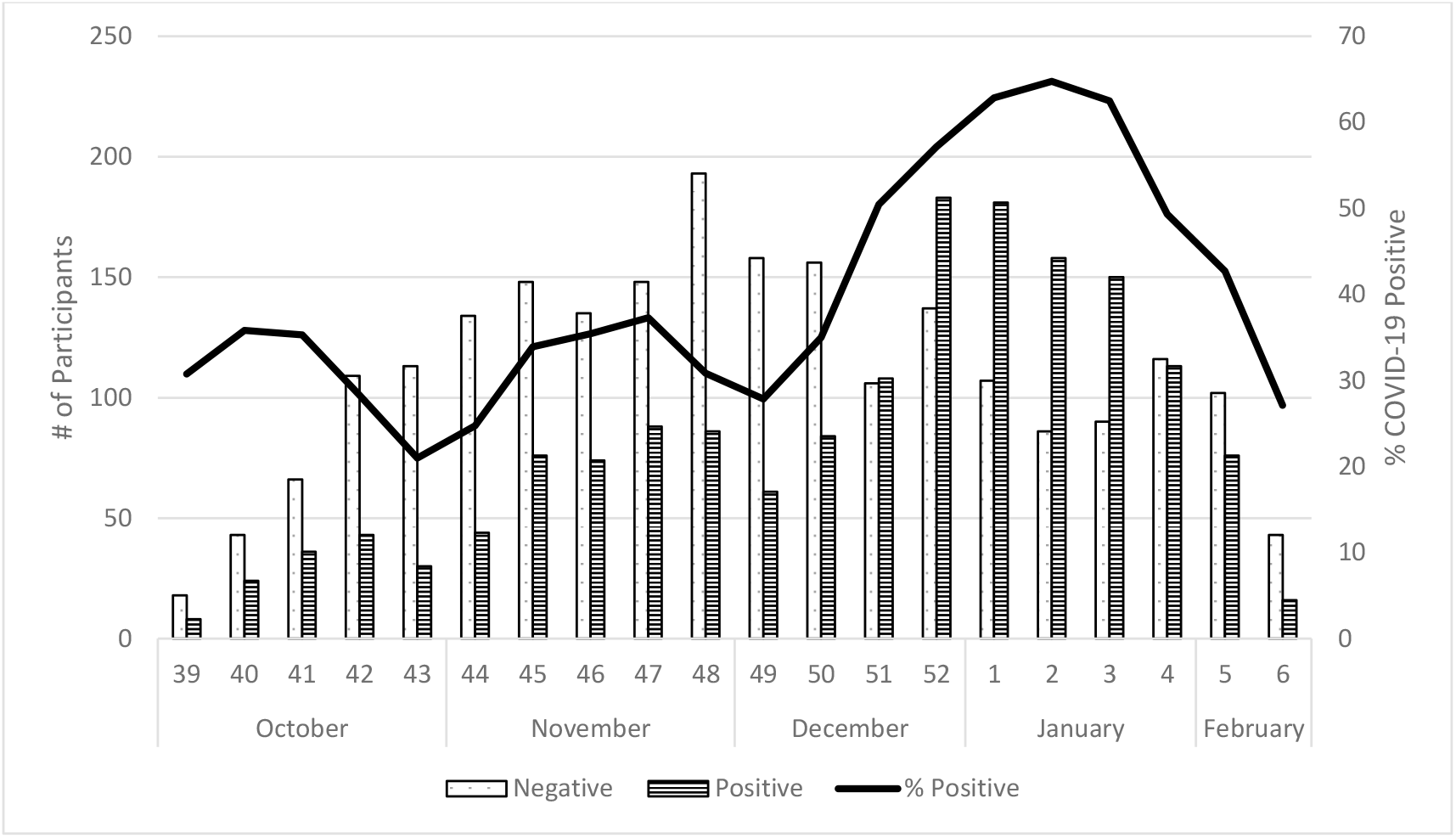
Number and Percent of SARS-CoV-2 Positive Enrolled Participants by Onset Week, US Flu VE Network, October 2021 – February 2022

Over the entire study period, participants who self-reported a high-risk exposure or reported fever were more likely to test positive (Table 1). Additionally, participants who were aged ≥65 years, identified as White non-Hispanic or other race non-Hispanic compared to Black non-Hispanic or Hispanic, self-reported a chronic medical condition, and did not self-report a fever were more likely to receive a 3^rd^ vaccine dose (Table 2). Among 2-dose recipients, the median interval between receipt of a 2^nd^ dose and illness onset date was 225 days (range:14 – 386); 13% and 87% had received a 2^nd^ mRNA vaccine dose 14-149 days or ≥150 days prior to illness onset, respectively (data not shown). The median interval between 3^rd^ dose receipt and illness onset was 53 days (range: 7 – 230) (data not shown).

**Table 1:** Characteristics of Symptomatic Adults Seeking Outpatient Medical Care or Clinical SARS-CoV-2 Testing by SARS-CoV-2 Status, US Flu VE Network, October 2021 – February 2022

|  | Negative SARS-CoV-2<br>Participants with CLI |  | Positive SARS-CoV-2<br>Participants with CLI |  | P-Value<sup>a</sup> |
| --- | --- | --- | --- | --- | --- |
|  | N | Col % | N | Col % |  |
| Total | 2208 | 100 | 1639 | 100 |  |
| Age Group, y |  |  |  |  | 0.06 |
| 18-49 | 1347 | 61 | 1027 | 63 |  |
| 50-64 | 544 | 25 | 419 | 26 |  |
| ≥65 | 317 | 14 | 193 | 12 |  |
| Site |  |  |  |  | <0.01 |
| California | 322 | 15 | 164 | 10 |  |
| Michigan | 166 | 8 | 188 | 11 |  |
| Pennsylvania | 311 | 14 | 320 | 20 |  |
| Tennessee | 269 | 12 | 186 | 11 |  |
| Texas | 262 | 12 | 159 | 10 |  |
| Washington | 296 | 13 | 110 | 7 |  |
| Wisconsin | 582 | 26 | 512 | 31 |  |
| Sex<sup>b</sup> |  |  |  |  | <0.01 |
| Female | 1485 | 67 | 1017 | 62 |  |
| Male | 720 | 33 | 618 | 38 |  |
| Race/Ethnicity<sup>c</sup> |  |  |  |  | <0.01 |
| Black, non-Hispanic | 87 | 4 | 119 | 7 |  |
| Hispanic | 221 | 10 | 134 | 8 |  |
| Other, non-Hispanic | 198 | 9 | 128 | 8 |  |
| White, non-Hispanic | 1676 | 77 | 1218 | 76 |  |
| Self-Reported Chronic Medical Condition<sup>d</sup> |  |  |  |  | 0.77 |
| No | 1514 | 70 | 1138 | 70 |  |
| Yes | 652 | 30 | 480 | 30 |  |
| High-Risk Exposure |  |  |  |  | <0.01 |
| No | 1273 | 58 | 706 | 43 |  |
| Yes | 935 | 42 | 933 | 57 |  |
| Self-Reported Prior Infection<sup>e</sup> |  |  |  |  | <0.01 |
| No | 1841 | 84 | 1450 | 89 |  |
| Yes (6 missing) | 350 | 16 | 179 | 11 | 0.12 |
| <3 months ago | 194 | 56 | 110 | 63 |  |
| ≥3 months ago | 154 | 44 | 65 | 37 |  |
| Product Among Vaccinated (2 and 3 Doses) |  |  |  |  |  |
| Moderna | 670 | 36 | 368 | 31 |  |
| Pfizer-BioNTech | 1165 | 62 | 784 | 67 |  |
| Combination | 42 | 2 | 20 | 2 |  |
| Fever<sup>f</sup> |  |  |  |  | <0.01 |
| No | 1233 | 56 | 649 | 40 |  |
| Yes | 950 | 44 | 968 | 60 |  |

Abbreviations: CLI – COVID-19-like illness
<sup>a</sup> P-value for chi-square statistic
<sup>b</sup> 7 participants missing data on sex
<sup>c</sup> 66 participants missing data on race and/or ethnicity
<sup>d</sup> 63 participants missing data on chronic medical condition
<sup>e</sup> 27 participants missing data on self-reported prior SARS-CoV-2 infection
<sup>f</sup> 47 participants missing data on presence of fever

**Table 2:** Characteristics of Symptomatic Adults Seeking Outpatient Medical Care or SARS-CoV-2 Clinical Testing by mRNA COVID-19 Verified Vaccination Status, US Flu VE Network, October 2021 – February 2022

|  | Unvaccinated |  | 2 Doses<br>(14-149 Days) |  | 2 Doses<br>(≥150 Days) |  | 3 Doses |  | P-Value<sup>a</sup> |
| --- | --- | --- | --- | --- | --- | --- | --- | --- | --- |
|  | N | Row % | N | Row % | N | Row % | N | Row % |  |
| Total | 798 | 21 | 256 | 7 | 1681 | 44 | 1112 | 29 |  |
| Age Group, y |  |  |  |  |  |  |  |  | <0.01 |
| 18-49 | 566 | 24 | 185 | 8 | 1099 | 46 | 524 | 22 |  |
| 50-64 | 181 | 19 | 63 | 7 | 399 | 41 | 320 | 33 |  |
| ≥65 | 51 | 10 | 8 | 2 | 183 | 36 | 268 | 53 |  |
| Site |  |  |  |  |  |  |  |  | <0.01 |
| California | 15 | 3 | 24 | 5 | 268 | 55 | 179 | 37 |  |
| Michigan | 50 | 14 | 15 | 4 | 183 | 52 | 106 | 30 |  |
| Pennsylvania | 208 | 33 | 29 | 5 | 292 | 46 | 102 | 16 |  |
| Tennessee | 56 | 12 | 44 | 10 | 194 | 43 | 161 | 35 |  |
| Texas | 130 | 31 | 49 | 12 | 179 | 43 | 63 | 15 |  |
| Washington | 12 | 3 | 23 | 6 | 210 | 52 | 161 | 40 |  |
| Wisconsin | 327 | 30 | 72 | 7 | 355 | 32 | 340 | 31 |  |
| Sex<sup>b</sup> |  |  |  |  |  |  |  |  | 0.09 |
| Female | 496 | 20 | 168 | 7 | 1086 | 43 | 752 | 30 |  |
| Male | 302 | 23 | 88 | 7 | 589 | 44 | 359 | 27 |  |
| Race/Ethnicity<sup>c</sup> |  |  |  |  |  |  |  |  | <0.01 |
| Black, non-Hispanic | 61 | 30 | 27 | 13 | 76 | 37 | 42 | 20 |  |
| Hispanic | 45 | 13 | 29 | 8 | 199 | 56 | 82 | 23 |  |
| Other, non-Hispanic | 32 | 10 | 16 | 5 | 170 | 52 | 108 | 33 |  |
| White, non-Hispanic | 646 | 22 | 180 | 6 | 1207 | 42 | 861 | 30 |  |
| Self-Reported Chronic Medical Condition<sup>d</sup> |  |  |  |  |  |  |  |  | <0.01 |
| No | 584 | 22 | 193 | 7 | 1190 | 45 | 685 | 26 |  |
| Yes | 205 | 18 | 61 | 5 | 461 | 41 | 405 | 36 |  |

Table 2 Continued:
|  | Unvaccinated |  | 2 Doses<br>(14-149 Days) |  | 2 Doses<br>(≥150 Days) |  | 3 Doses |  | P-Value<sup>a</sup> |
| --- | --- | --- | --- | --- | --- | --- | --- | --- | --- |
|  | N | Row % | N | Row % | N | Row % | N | Row % |  |
| High-Risk Exposure |  |  |  |  |  |  |  |  | <0.01 |
| No | 406 | 21 | 134 | 7 | 916 | 46 | 523 | 26 |  |
| Yes | 392 | 21 | 122 | 7 | 765 | 41 | 589 | 32 |  |
| Self-reported Prior Infection<sup>e</sup> |  |  |  |  |  |  |  |  | <0.01 |
| No | 626 | 19 | 198 | 6 | 1466 | 45 | 1001 | 30 |  |
| Yes (6 missing) | 169 | 32 | 55 | 10 | 200 | 38 | 105 | 20 | <0.01 |
| <3 months ago | 123 | 40 | 25 | 8 | 96 | 32 | 60 | 20 |  |
| ≥3 months ago | 45 | 21 | 29 | 13 | 101 | 46 | 44 | 20 |  |
| Product Among Vaccinated (2 and 3 Doses) |  |  |  |  |  |  |  |  |  |
| Moderna | . | . | 64 | 6 | 636 | 61 | 338 | 33 |  |
| Pfizer-BioNTech | . | . | 190 | 10 | 1045 | 54 | 714 | 37 |  |
| Combination | . | . | 2 | 3 | 0 | 0 | 60 | 97 |  |
| Fever<sup>f</sup> |  |  |  |  |  |  |  |  | <0.01 |
| No | 325 | 17 | 120 | 6 | 795 | 42 | 642 | 34 |  |
| Yes | 468 | 24 | 135 | 7 | 862 | 45 | 453 | 24 |  |

<sup>a</sup> P-value for chi-square statistic<sup>b</sup> 7 participants missing data on sex<sup>c</sup> 66 participants missing data on race and/or ethnicity<sup>d</sup> 63 participants missing data on chronic medical condition<sup>e</sup> 27 participants missing data on self-reported prior SARS-CoV-2 infection<sup>f</sup> 47 participants missing data on presence of fever

### Study Periods by Variant Predominance

Sequencing results from 272 US Flu VE Network participants with onset dates between November 9, 2021, and January 9, 2022, demonstrated distinct periods of Delta versus Omicron circulation with co-circulation of both variants during December 10 – 19, 2021 (Figure 2). Overall, 45% of sequenced specimens were Delta. The first Omicron variant in the network was detected on December 10, 2021, and it became the consistently predominant variant (>50% of sequenced viruses) by December 15, 2021, with few viruses in early January 2022 still being identified as Delta.

**Figure 2:**
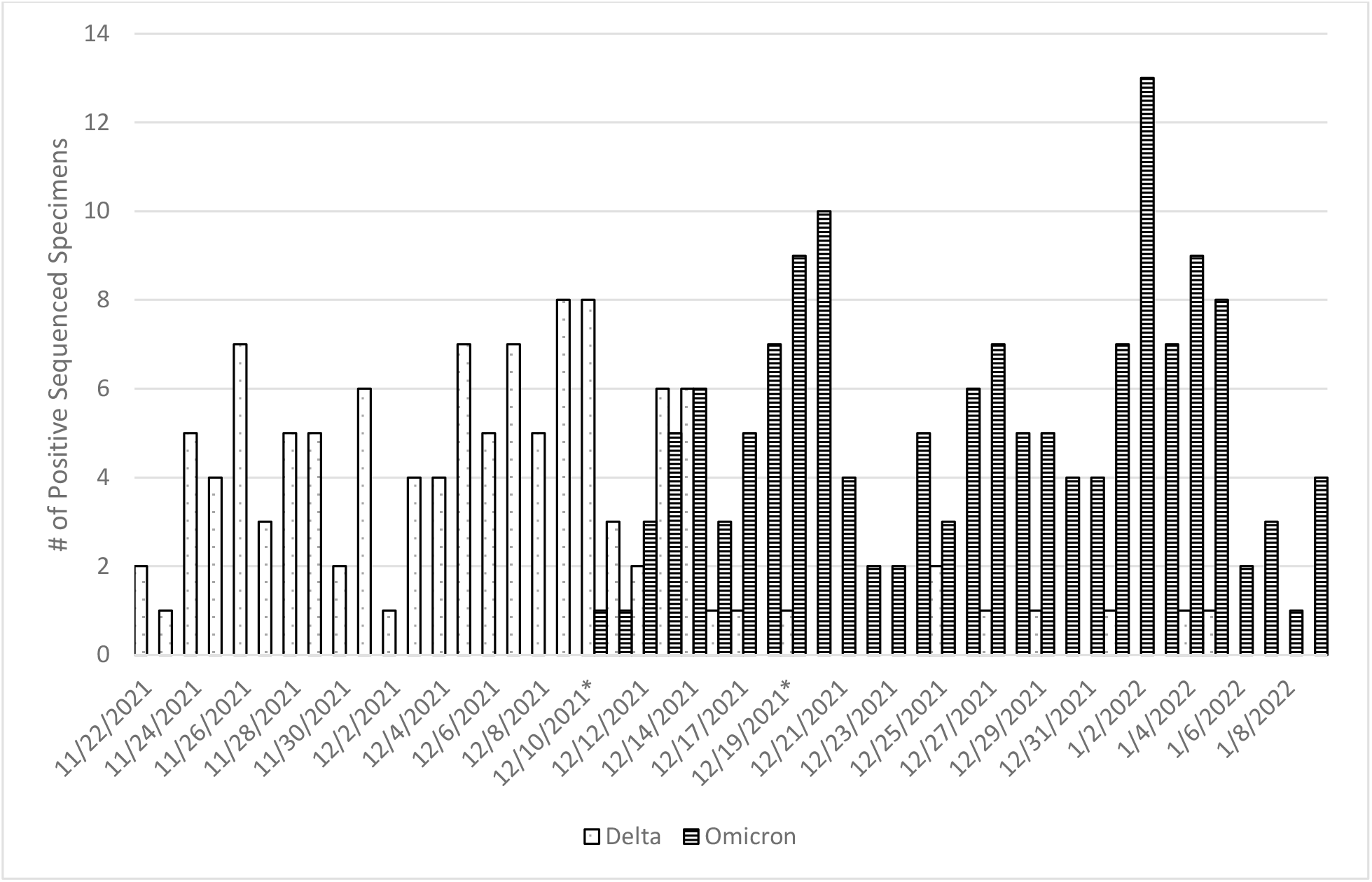
SARS-CoV-2 Variant Virus Distribution among Subset of Symptomatic SARS-CoV-2 Positive Participants by Onset Date, November 2021 – January 2022 *Variant transition period in the US Flu VE Network: December 10 – December 19, 202

### Vaccine Effectiveness

During the Delta period, adjusted VE against symptomatic illness in outpatient settings was 63% (95% CI: 51% to 72%) among mRNA 2-dose recipients and 96% (95% CI: 93% to 98%) for 3-dose recipients (Table 3). During the Omicron period, adjusted VE was 21% (95% CI: -6% to 41%) among 2-dose recipients and 62% (95% CI: 48% to 72%) among 3-dose recipients. During the Delta period, VE among participants who received their 2^nd^ dose 14-149 days before illness onset was 89% (95% CI: 78% to 94%) compared to 58% (95% CI: 44% to 68%) among those who received their 2^nd^ dose ≥150 days before illness onset (Table 3). During the Omicron period, VE among those who received their 2^nd^ dose 14-149 days before illness onset was 45% (14% to 66%) and among those who received their 2^nd^ dose ≥150 days before illness onset was 11% (−21% to 35%). Excluding 3-dose recipients who received the 3^rd^ dose <150 days after the 2^nd^ dose (n=35) did not change 3-dose VE estimates (data not shown).

**Table 3:** 2- and 3-Dose Vaccine Effectiveness during SARS-CoV-2 Delta Variant or Omicron Variant Associated Symptomatic COVID-19 Illness among Adults Seeking Outpatient Medical Care or SARS-CoV-2 Clinical Testing, US Flu VE Network, October 2021 – February 2022

|  | SARS-CoV-2 Positive |  | SARS-CoV-2 Negative |  | Unadjusted VE |  | Adjusted<sup>a</sup> VE |  |
| --- | --- | --- | --- | --- | --- | --- | --- | --- |
|  | Vaccinated/ Total | (%) | Vaccinated/ Total | (%) | VE | (95% CI) | VE | (95% CI) |
| **Overall** |  |  |  |  |  |  |  |  |
| 2-Dose | 822/1289 | (64) | 1115/1446 | (77) | 48 | (38 to 56) | 48 | (37 to 57) |
| 3-Dose | 350/817 | (43) | 762/1093 | (70) | 67 | (61 to 73) | 78 | (72 to 83) |
| **Delta<sup>b</sup>** |  |  |  |  |  |  |  |  |
| 2-Dose | 327/552 | (59) | 763/942 | (81) | 66 | (57 to 73) | 63 | (51 to 72) |
| 14-149 Days | 14/239 | (6) | 106/285 | (37) | 89 | (81 to 94) | 89 | (78 to 94) |
| ≥150 Days | 313/538 | (58) | 657/836 | (79) | 62 | (52 to 70) | 58 | (44 to 68) |
| 3-Dose | 22/247 | (9) | 259/438 | (59) | 93 | (89 to 96) | 96 | (93 to 98) |
| **Omicron<sup>b</sup>** |  |  |  |  |  |  |  |  |
| 2-Dose | 464/684 | (68) | 257/380 | (68) | 0 | (-32 to 23) | 21 | (-6 to 41) |
| 14-149 Days | 69/289 | (24) | 53/176 | (30) | 27 | (-11 to 52) | 45 | (14 to 66) |
| ≥150 Days | 395/615 | (64) | 204/327 | (62) | -8 | (-43 to 18) | 11 | (-21 to 35) |
| 3-Dose | 322/542 | (59) | 408/531 | (77) | 56 | (43 to 66) | 62 | (48 to 72) |

<sup>a</sup>Logistic regression model adjusted for age, site, illness onset week, and prior infection status.
<sup>b</sup>Totals in variant-specific periods may not add up to overall total as a transition period was included in the overall estimates but removed in the variant-specific periods

### Vaccine Effectiveness by Subgroup

Self-reported high-risk exposure status, self-reported presence of a chronic medical condition, self-reported prior laboratory-confirmed SARS-CoV-2 infection, longer interval from illness onset to respiratory specimen collection, and self-reported presence of fever with cough or shortness of breath did not change 3-dose VE during the Delta variant predominant period (Table 4). However, during the period when the Omicron variant predominated, 3-dose VE point estimates tended to be lower but with overlapping confidence intervals among those who had a high-risk exposure, a chronic medical condition, a prior SARS-CoV-2 infection, or CLI that included fever. During the Delta period, 4% of cases and 14% of controls had prior infection compared to the Omicron period when 15% of cases and 20% of controls had prior infection.

**Table 4:** Results of Subgroup Analyses of 3-Dose Vaccine Effectiveness against Delta and Omicron Variant Related Symptomatic COVID-19 Illness

|  | SARS-CoV-2 Positive |  | SARS-CoV-2 Negative |  | Unadjusted VE |  | Adjusted<sup>a</sup> VE |  |
| --- | --- | --- | --- | --- | --- | --- | --- | --- |
|  | Vaccinated/ Total | (%) | Vaccinated/ Total | (%) | VE | (95% CI) | VE | (95% CI) |
| **Delta** |  |  |  |  |  |  |  |  |
| High-Risk Exposure |  |  |  |  |  |  |  |  |
| No | 9/107 | (8) | 145/261 | (56) | 93 | (85 to 96) | 98 | (94 to 99) |
| Yes | 13/140 | (9) | 114/177 | (64) | 94 | (89 to 97) | 96 | (92 to 98) |
| Chronic Medical Condition |  |  |  |  |  |  |  |  |
| No | 7/161 | (4) | 134/259 | (52) | 96 | (91 to 98) | 98 | (94 to 99) |
| Yes | 15/84 | (18) | 120/168 | (71) | 91 | (83 to 95) | 95 | (87 to 98) |
| Prior Infection |  |  |  |  |  |  |  |  |
| No | 20/228 | (9) | 239/378 | (63) | 94 | (91 to 97) | 97 | (95 to 99) |
| Yes | 1/17 | (6) | 20/60 | (33) | 87 | (-1 to 98) | 79 | (-81 to 98) |
| Days from Illness Onset to Respiratory Specimen Collection |  |  |  |  |  |  |  |  |
| 0-2 Days | 17/206 | (8) | 212/355 | (60) | 94 | (90 to 96) | 97 | (94 to 99) |
| 3-10 Days | 5/41 | (12) | 47/83 | (57) | 89 | (70 to 96) | 93 | (70 to 98) |
| Fever + Cough/Shortness of Breath |  |  |  |  |  |  |  |  |
| No | 9/72 | (13) | 131/195 | (67) | 93 | (85 to 97) | 95 | (86 to 98) |
| Yes | 13/175 | (7) | 128/243 | (53) | 93 | (87 to 96) | 97 | (94 to 99) |
| **Omicron** |  |  |  |  |  |  |  |  |
| High-Risk Exposure |  |  |  |  |  |  |  |  |
| No | 111/218 | (51) | 199/253 | (79) | 72 | (58 to 81) | 76 | (61 to 86) |
| Yes | 211/324 | (65) | 209/278 | (75) | 38 | (12 to 57) | 49 | (23 to 66) |
| Chronic Medical Condition |  |  |  |  |  |  |  |  |
| No | 201/378 | (53) | 266/360 | (74) | 60 | (45 to 71) | 66 | (50 to 76) |
| Yes | 117/159 | (74) | 131/160 | (82) | 38 | (-5 to 64) | 45 | (-4 to 71) |
| Prior Infection |  |  |  |  |  |  |  |  |
| No | 291/460 | (63) | 356/428 | (83) | 65 | (52 to 75) | 64 | (49 to 75) |
| Yes | 29/79 | (37) | 49/99 | (49) | 41 | (-8 to 68) | 52 | (-1 to 77) |
| Days from Illness Onset to Respiratory Specimen Collection |  |  |  |  |  |  |  |  |
| 0-2 Days | 295/487 | (61) | 360/472 | (76) | 52 | (37 to 64) | 60 | (44 to 71) |
| 3-10 Days | 27/55 | (49) | 48/59 | (81) | 78 | (49 to 90) | 72 | (18 to 91) |
| Fever + Cough/Shortness of Breath |  |  |  |  |  |  |  |  |
| No | 142/215 | (66) | 221/261 | (85) | 65 | (45 to 77) | 68 | (47 to 81) |
| Yes | 180/327 | (55) | 187/270 | (69) | 46 | (24 to 61) | 55 | (31 to 70) |

<sup>a</sup>Logistic regression model adjusted for age, site, illness onset week, and prior infection status.

## Discussion

This investigation adds to early evidence of effectiveness of a 3^rd^ mRNA vaccine dose against laboratory-confirmed SARS-CoV-2 infection among adults seeking outpatient care and clinical testing for CLI symptoms during the pandemic wave predominated by the Omicron variant [3, 11-14]. However, 3-dose effectiveness among adults was lower during the Omicron predominant period than during the pandemic wave associated with the Delta variant. Similar to analyses of large electronic medical record databases or data from SARS-CoV-2 testing sites, 3-dose VE in this analysis was higher against Delta than against Omicron-related illness [3, 11-12].

Findings from the US Flu VE Network are also consistent with higher estimates of 2-dose VE when the 2^nd^ dose was given less than 5 months before current illness onset compared to at least 5 months or more before illness onset [3, 5]. Waning effectiveness against SARS-CoV-2 Delta variant virus infection or associated outpatient illness was also observed 5 to 6 months after receipt of the 2^nd^ mRNA vaccine dose in other countries using multiple study designs [15-20]. However, among US Flu VE Network participants, 2-dose mRNA VE point estimate against outpatient illness associated with the Delta variant among those who received their 2^nd^ dose at least 5 months or more before illness onset remained higher than 2-dose VE against Omicron among those who received their 2^nd^ dose <5 months before illness onset, with overlapping 95% confidence intervals. These results suggest that updates to COVID-19 vaccine formulations or additional booster doses may be needed to improve protection against future SARS-CoV-2 variant viruses.

Active enrollment of study participants in the US Flu VE Network provided additional information to evaluate differences in 3-dose mRNA VE according to participants’ symptoms, reported history of past laboratory-confirmed SARS-CoV-2 infection, high-risk exposure, and presence of underlying medical conditions. First, among generally healthy outpatients with symptomatic illness enrolled in the US Flu VE Network, 3-dose VE point estimates during the Omicron period tended to be lower among participants reporting underlying medical conditions compared to point estimates among participants without underlying conditions. Presence of underlying medical conditions, especially immunosuppressive conditions, have been associated with decreased mRNA VE against severe outcomes including COVID-19 related hospitalizations [3, 21-28], and provided the basis for the recommendation of a 3^rd^ primary mRNA vaccine dose [1]. Second, participants who reported a high-risk exposure in the 14 days before illness onset demonstrated lower 3-dose VE during the Omicron predominant period compared with overall VE during this time. These results are consistent with previous studies, including an analysis of data from the US Flu VE Network during the Delta-predominant period [6, 29, 30]. Third, the proportion of participants reporting previous laboratory confirmed SARS-CoV-2 infection was higher when the Omicron variant predominated than when the Delta variant predominated. However, we were unable to evaluate the impact of time since prior infection on VE due to small sample sizes. In contrast, prior studies have demonstrated increased protection among persons with prior SARS-CoV-2 infection history [31,32].

This investigation is subject to at least six limitations. First, small sample sizes limited our ability to evaluate VE by certain subgroups. Differences between 2- and 3-dose mRNA VE by vaccine product, age group, and underlying medical conditions have been reported from studies including larger numbers of patients or medical encounters [3, 5, 11-15]. Second, adolescents and children were not included in this analysis due to lower proportion of enrollment than in typical influenza seasons and lower percent vaccinated. Third, because of recent authorization of a booster dose for adults, waning of 3-dose VE could not be assessed. Waning effectiveness of a booster dose against COVID-19 associated emergency department or urgent care visits has been reported elsewhere, though the study population may have differed to a certain extent from that of the US Flu VE Network [5]. Fourth, with active enrollment, persons consenting to participate may differ from all patients in ways that may affect VE estimates, such as different healthcare-seeking behaviors among vaccinated and unvaccinated persons [10]. Vaccinated SARS-CoV-2 positive participants may have been more likely than unvaccinated positive participants to participate in this study. Fifth, Delta versus Omicron misclassification among the subset of infections without sequencing results is possible. Finally, increased use of at-home testing may result in changes in healthcare seeking behavior and potential biases for VE studies, which requires further examination.

VE studies that rely on active enrollment of patients meeting clinical criteria for acute respiratory illness may contribute to ongoing monitoring of effectiveness of current and future COVID-19 vaccines [8]. Studies in this outpatient setting also contribute to understanding vaccine protection against a spectrum of illness, adding effectiveness against symptomatic illness in outpatient settings to published inpatient, emergency department, and urgent care estimates for moderately severe and severe COVID-19. Systematic testing of outpatients presenting with CLI has the potential to identify SARS-CoV-2 positive cases and collect vaccination histories for VE estimates that may not be available from analyses of electronic medical records, especially as SARS-CoV-2 testing for persons with symptomatic illness becomes less frequent [33]. As SARS-CoV-2 viruses evolve and COVID-19 may continue to cause influxes of respiratory illness, systematic testing for respiratory illnesses including COVID-19 and influenza will be important to evaluate effectiveness of COVID-19 vaccines and immunization schedules.

## Data Availability

De-identified data produced in the present study may be available upon reasonable request to the authors.

## Acknowledgements

Alexander Arroliga, Madhava Beeram, Kayan Dunnigan, Jason Ettlinger, Ashley Graves, Eric Hoffman, Muralidhar Jatla, Amanda McKillop, Kempapura Murthy, Manohar Mutnal, Elisa Priest, Chandni Raiyani, Arundhati Rao, Lydia Requenez, Natalie Settele, Michael Smith, Keith Stone, Jennifer Thomas, Marcus Volz, Kimberly Walker, Martha Zayed, Baylor Scott & White Health, Temple, Texas and Texas A&M University College of Medicine, Temple, Texas; Ekow Annan, Peter Daley, Krista Kniss, Angiezel Merced-Morales, Influenza Division, CDC, Atlanta, GA; Elmer Ayala, Britta Amundsen, Michael Aragones, Raul Calderon, Vennis Hong, Gabriela Jimenez, Jeniffer Kim, Jen Ku, Bruno Lewin, Ashley McDaniel, Alexandria Reyes, Sally Shaw, Harp Takhar, Alicia Torres, Pasadena Medical Office Urgent Care Staff, Kaiser Permanente Southern California, Pasadena, California; Rachael Burganowski, Erika Kiniry, Kathryn A. Moser, Matt Nguyen, Suzie Park, Stacie Wellwood, Brianna Wickersham, Kaiser Permanente Washington Health Research Institute, Seattle, Washington; Juan Alvarado-Batres, Saydee Benz, Hannah Berger, Adam Bissonnette, Joshua Blake, Krystal Boese, Emily Botten, Jarod Boyer, Michaela Braun, Brianna Breu, Gina Burbey, Caleb Cravillion, Christian Delgadillo, Amber Donnerbauer, Tim Dziedzic, Joseph Eddy, Heather Edgren, Alex Ermeling, Kelsey Ewert, Connie Fehrenbach, Rachel Fernandez, Wayne Frome, Sherri Guzinski, Linda Heeren, David Herda, Mitchell Hertel, Garrett Heuer, Erin Higdon, Lynn Ivacic, Lee Jepsen, Steve Kaiser, Julie Karl, Bailey Keffer, Jennifer King, Tamara Kronenwetter Koepel, Stephanie Kohl, Sarah Kohn, Diane Kohnhorst, Erik Kronholm, Thao Le, Alaura Lemieux, Carrie Marcis, Megan Maronde, Isaac McCready, Karen McGreevey, Jennifer Meece, Nidhi Mehta, Daniel Miesbauer, Vicki Moon, Jennifer Moran, Collin Nikolai, Brooke Olson, Jeremy Olstadt, Lisa Ott, Nan Pan, Cory Pike, DeeAnn Polacek, Martha Presson, Nicole Price, Christopher Rayburn, Chris Reardon, Miriah Rotar, Carla Rottscheit, Jacklyn Salzwedel, Juan Saucedo, Kelly Scheffen, Charity Schug, Kristin Seyfert, Ram Shrestha, Alexander Slenczka, Elisha Stefanski, Melissa Strupp, Megan Tichenor, Lyndsay Watkins, Anna Zachow, Ben Zimmerman, Marshfield Clinic Research Institute, Marshfield, Wisconsin; Sarah Bauer, Kim Beney, Caroline K. Cheng, Nahla Faraj, Amy Getz, Michelle Grissom, Michelle Groesbeck, Samantha Harrison, Kristen Henson, Kim Jermanus, Emileigh Johnson, Anne Kaniclides, Armanda Kimberly, Lois E. Lamerato, Adam Lauring, Regina Lehmann-Wandell, E. J. McSpadden, Louis Nabors, Rachel Truscon, University of Michigan, Ann Arbor, Michigan and Henry Ford Health System, Detroit, Michigan; G.K. Balasubramani, Todd Bear, Jared Bobeck, Erin Bowser, Karen Clarke, Lloyd G. Clarke, Klancie Dauer, Chris Deluca, Blair Dierks, Linda Haynes, Robert Hickey, Monika Johnson, Andrea Jonsson, Nancy Luosang, Leah McKown, Alanna Peterson, Demetria Phaturos, Andrew Rectenwald, Theresa M. Sax, Miles Stiegler, Michael Susick, Joe Suyama, Louise Taylor, Sara Walters, Alexandra Weissman, John V. Williams, University of Pittsburgh Schools of the Health Sciences and University of Pittsburgh Medical Center, Pittsburgh, Pennsylvania; Marcia Blair, Juliana Carter, Jim Chappell, Emma Copen, Meredith Denney, Kellie Graes, Natasha Halasa, Chris Lindsell, Zhouwen Liu, Stephanie Longmire, Rendie McHenry, Laura Short, His-Nien Tan, Denise Vargas, Jesse Wrenn, Dayna Wyatt, Yuwei Zhu, Vanderbilt University Medical Center, Nashville, Tennessee; Strain Surveillance and Emerging Variant Bioinformatics Working Group.

## Footnotes

1 See 45 C.F.R. part 46; 21 C.F.R part 56

